# Quality assessment of data for decentralised antiretroviral therapy referrals and laboratory results in the South African national electronic HIV management register TIER.Net

**DOI:** 10.1101/2025.11.11.25340044

**Authors:** Lisanthini Naidu, Johan van der Molen, Vishen Jugathpal, Yukteshwar Sookrajh, Thokozani Khubone, Lungile Hobe, Thulani Ngwenya, Kwena Tlhaku, Sharana Mahomed, Nigel Garrett, Jennifer Anne Brown, Jienchi Dorward, Lara Lewis

## Abstract

Three Interlinked Electronic Register (TIER.Net) is South Africa’s national electronic HIV patient database, used to monitor antiretroviral therapy (ART) delivery and laboratory results. However, few published evaluations have quantified TIER.Net data quality relative to national sources. We aimed to evaluate how well decentralised ART referral and laboratory result data are captured in TIER.Net.

We conducted a retrospective analysis comparing TIER.Net to national electronic health systems. For decentralised ART, we used de-identified data from 56 clinics in eThekwini (2020-2023) and compared the annual number of TIER.Net decentralised ART referrals to ART prescriptions in the Synchronised National Communication in Health (SyNCH) database. For laboratory data, we used de-identified records from 103 clinics in KwaZulu-Natal (2015-2022) and compared the annual number of TIER.Net viral load (VL) and CD4 tests with the number in National Health Laboratory Service (NHLS). The proportion of SyNCH decentralised ART prescriptions and NHLS VL and CD4 counts captured in TIER.Net were calculated by clinic, and trends were assessed using linear mixed-effects models (LMMs).

The median proportion of SyNCH decentralised ART prescriptions captured in TIER.Net was 104.4% (IQR: 99.85-115.1*%*) in 2020 and 102.44% (IQR: 100.5-104.5%) in 2023. The LMM estimated an annual decrease of 2.8% (95% CI: -4.8; -0.9%). The median proportion of NHLS VLs captured in TIER.Net was 85.7% (IQR: 70.0-97.9%) in 2015 and 99.1% (IQR: 94.5-102.5%) in 2022. The LMM estimated an annual increase of 1.8% (95% CI: 1.4-2.1%). The median proportion of NHLS CD4s captured in TIER.Net was 74.3% (IQR: 63.9-85.4%) in 2015 and 80.1% (IQR: 68.4-89.1%) in 2022. The LMM estimated no statistically significant trend over time (-0.08%, 95% CI: -0.6; 0.4%).

Reassuringly, capture of TIER.Net for decentralised ART and VL data has improved to near 100%, but CD4 count capture remains sub-optimal, highlighting strengths and limitations of conducting analyses with this critical HIV programme database.

## Introduction

The Three Interlinked Electronic Register (TIER.Net) is a paper-to-digital electronic register used to record demographic and clinical data for individuals receiving antiretroviral therapy (ART) in the South African public sector [1]. TIER.Net data capturers use paper-based medical records to manually enter data on decentralised ART delivery through the Centralized Chronic Medicines Dispensing and Distribution (CCMDD) programme and laboratory test results including viral load (VL) and CD4 counts. Data from TIER.Net is collated at clinic, district, provincial, and national levels to support the evaluation of the HIV programme and to support clinical management and policy decisions [1-3]. However, few published evaluations have quantified the quality of TIER.Net data relative to other national data sources that record CCMDD ART prescriptions, VL and CD4 count tests.

In South Africa, decentralised ART delivery for clinically stable people living with HIV has expanded rapidly through the CCMDD programme, which enables patients to collect their medicines from designated pick-up points outside of healthcare facilities [4]. By 2024, over 5.3 million patients had been registered on the CCMDD programme with approximately 2.9 million patients actively collecting ART through the programme [5]. For patients receiving an ART prescription through CCMDD, the corresponding clinic visit should be manually recorded in TIER.Net as a CCMDD referral. Accurate capture of these referrals in TIER.Net is critical to ensure that patients collecting ART outside of the clinic are not erroneously classified as lost to follow-up. CCMDD prescriptions, including ART prescriptions, are processed through the Synchronised National Communication in Health (SyNCH) [6, 7]. Therefore, comparison of SyNCH data and TIER.Net data can affirm the completeness and reliability of CCMDD referral records within TIER.Net.

The National Health Laboratory Service (NHLS) conducts all public sector VL and CD4 count testing and has a national repository for laboratory data from the public sector [8]. Therefore, comparison of NHLS and TIER.Net data can affirm the reliability of VL and CD4 count records captured within TIER.Net.

Understanding the quality of data within TIER.Net is increasingly critical as South Africa scales up differentiated service delivery models such as the CCMDD programme. With rising patient enrolment in CCMDD and a growing emphasis on client-centred care amidst funding constraints, accurate and reliable data is essential for monitoring programme performance and informing policy decisions. TIER.Net provides a comprehensive dataset, including sociodemographic, clinical, and laboratory variables necessary for robust evaluations of CCMDD’s impact on treatment outcomes [4, 9]. Key indicators such as VL, which reflects the final “95” in the UNAIDS 95-95-95 targets [10], and CD4 count, used to classify AIDS progression and advanced HIV disease (AHD) [11], are central to programmatic assessments. Furthermore, TIER.Net data has supported numerous peer-reviewed publications evaluating various aspects of the national HIV response [3, 12-14]. A deeper understanding of the completeness and reliability of this data will help identify and mitigate potential biases in current and future analyses.

We aimed to assess the quality of data captured in TIER.Net by comparing the total number of CCMDD referrals in TIER.Net with the corresponding CCMDD ART prescriptions fulfilled in the SyNCH database, as well as the total number of VL and CD4 tests recorded in TIER.Net with the corresponding results in the NHLS database.

## Methods

### Study design and setting

We conducted a retrospective analysis using de-identified, routinely collected data from the KwaZulu-Natal province, South Africa. To compare decentralised ART referral data, we included clinic-level TIER.Net and SyNCH data from 56 public clinics in the eThekwini Municipality. To compare VL and CD4 count data, we used clinic-level TIER.Net and NHLS data from 103 clinics managed by eThekwini Municipality (53 clinics), uMgungudlovu District (37 clinics) and uMkhanyakude District (13 clinics).

In 2022, KwaZulu-Natal was among the provinces with the highest HIV prevalence (16.0%) [15]. The CCMDD programme was initially introduced in KwaZulu-Natal to support ART roll-out [12]. The programme contracts pharmacy service providers to dispense and deliver ART and noncommunicable disease medicine parcels at pick-up points, including external pick-up points (private sector community pharmacies or general practitioners), adherence clubs, and internal pick-up points within the clinic that allow for ART collection without a nurse consultation [7]. The standard prescription length is 6-months, however, 12-month prescriptions were permitted from May 2020 to September 2021 in response to the COVID-19 pandemic [9].

Based on the national ART guidelines applicable during the time of this analysis, HIV VL testing was advised at six and twelve months after starting ART, and then annually, with viraemia triggering adherence counselling and repeat VL testing after three months [16]. In South Africa, the ‘Universal Test and Treat’ (UTT) strategy was rolled out in May 2016 which advises ART initiation to individuals who test positive for HIV irrespective of their CD4 count and clinical staging [17]. CD4 cell count testing is advised at and 10 months after ART initiation, upon re-initiation after disengagement from care, detection of viraemia, or clinical indication, and six-monthly while viraemic or as long as the previous CD4 cell count was ≤200 cells/µL [16]. CD4 counts are a crucial predictor of disease progression, particularly for AHD [11].

### Participants

We included all people living with HIV receiving ART through the CCMDD programme at participating clinics in eThekwini Municipality between 1 January 2020 and 31 December 2023, and all HIV VL and CD4 test results recorded in TIER.Net and the NHLS data warehouse from participating clinics between 1 January 2015 and 31 December 2022.

### Data sources and data management

TIER.Net, NHLS and SyNCH data were accessed on 19 March 2024, 30 Jan 2025 and 29 May 2025, respectively. TIER.Net and NHLS included individual de-identified patient level data, while SyNCH included aggregated clinic-level data. TIER.Net CCMDD ART referrals and SyNCH CCMDD ART prescriptions were categorised by pick-up point (external, internal and adherence club) by year and totalled to the clinic-level by year. TIER.Net and NHLS VL and CD4 count tests were totalled by thresholds by year and totalled to clinic-level by year.

### Variables

For the CCMDD analysis, the primary endpoint was the proportion of SyNCH CCMDD ART prescriptions captured in TIER.Net per year. The secondary endpoints were the proportion of SyNCH CCMDD ART prescriptions captured in TIER.Net per year by pick-up point (external, internal and adherence club). For the laboratory analysis, the primary endpoints were the proportion of NHLS VL and CD4 cell count results captured in TIER.Net per year. The secondary endpoints were proportion of NHLS VL and CD4 cell count results in TIER.Net per year by threshold (VL: <50, between 50 and 1000, and ≥ 1000 copies/mL; CD4: < 200 and ≥ 200 cells/µL). Clinic size was measured annually and defined as the number of individuals who visited the clinic at least once in a year.

### Statistical analysis

To describe the clinics used in the analysis, the median and interquartile range (IQR) clinic size was illustrated by year and district. We created figures showing yearly totals of SyNCH CCMDD ART prescriptions and TIER.Net CCMDD ART referral visits overall and by pick-up point. We also created figures showing yearly totals of NHLS and TIER.Net VL and CD4 counts overall and by threshold. We divided the total number of TIER.Net CCMDD ART referrals by the corresponding total number of SyNCH CCMDD ART prescriptions to obtain proportions by clinic, and similarly we divided the total number of VL and CD4 count results in TIER.Net by the corresponding numbers in NHLS to obtain proportions by clinic. We calculated and plotted the median and IQR of the clinic-level proportions per year. Outliers were defined as clinic-level proportions which are more than 3 standard deviations away from the mean. Linear mixed models (LMM) were used to analyse clinic-level proportions for the three outcomes (CCMDD prescriptions, VL and CD4 counts), with fixed effects for year and district and clinic-specific intercepts. For the laboratory outcomes, we conducted a sensitivity analysis to investigate whether trends over time differed by district. Specifically, we used a likelihood ratio test to assess whether the inclusion of an interaction term between year and district improved model fit. Data was analysed using R version 4.1.1, and we used a significance level of 0.05.

### Ethical approval

This work was approved by University of KwaZulu-Natal Biomedical Research Ethics Committee (BE646/17), the KwaZulu-Natal Department of Health’s Provincial Health Research Ethics Committee (KZ_201807_021), NHLS (PR2346201) and the eThekwini Municipality Health Unit, with a waiver for informed consent for analysis of de-identified, routinely collected data.

## Results

### Description of included clinics

We included 58 clinics (51 clinics contributing to the CCMDD and VL and CD4 dataset, 5 clinics contributing to the CCMDD dataset only and 2 clinics contributing to the VL and CD4 dataset only) in eThekwini, with a median size of 1862 in 2015 and 3412 in 2023, 37 clinics from uMgungudlovu with a median size of 1214 in 2015 and 2531 in 2023, and 13 clinics in uMkhanyakude, with a median size of 1083 in 2015 and 1488 in 2023. (Fig 1). We excluded 4 clinics due to outlying CCMDD or lab data.

**Fig 1.**
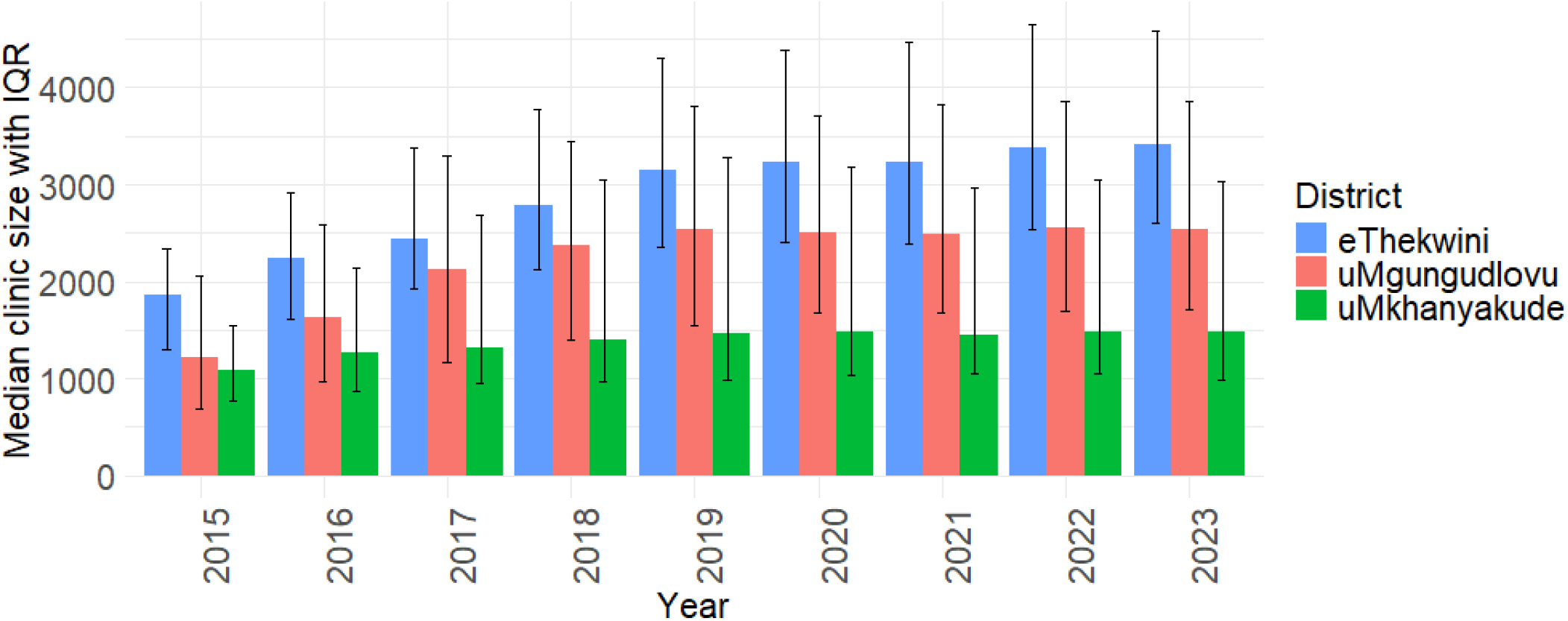
Median clinic size by district and year, with bars indicating interquartile ranges (IQRs).

### CCMDD data

Between 2020-2023, there were 741,211 CCMDD referral visits in TIER.Net and 715,063 ART CCMDD prescriptions in SyNCH. CCMDD referral visits in TIER.Net increased from 162,818 in 2020 to 238,182 in 2023 while ART CCMDD prescriptions in SyNCH increased from 147,281 in 2020 to 233,214 in 2023 (Fig 2A). In 2020, the median (IQR) clinic-level proportion of total ART CCMDD prescriptions in SyNCH captured in TIER.Net as CCMDD referral visits was 104.40% (99.85 to 115.13%), which decreased to 102.44% (100.45 to 104.54%) (Fig 2B) by 2023. The LMM estimated an annual decrease of 2.84% (95% CI: - 4.79 to -0.90%) in the proportion of SyNCH CCMDD ART prescriptions captured in TIER.Net.

**Fig 2.**
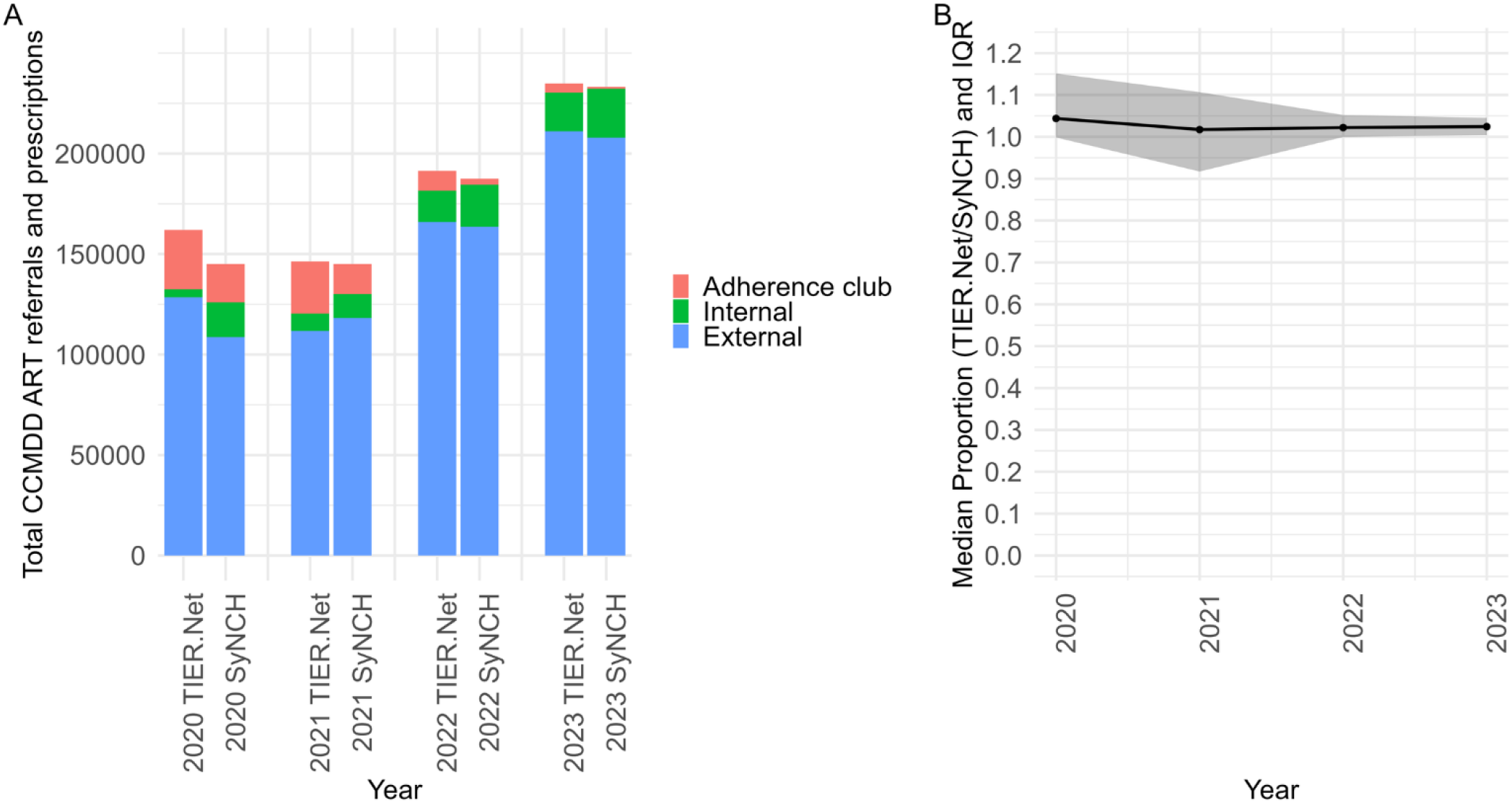
(A) Total number of CCMDD ART referrals (TIER.Net) and prescriptions (SyNCH) by pick-up point type and year. (B) Median clinic-level proportion of CCMDD prescriptions with recorded CCMDD referrals (TIER.Net/SyNCH) by year, with shading indicating the IQR. ART: antiretroviral therapy; CCMDD: Centralized Chronic Medicines Dispensing and Distribution; IQR: interquartile range; SyNCH: Synchronised National Communication in Health; TIER.Net: Three Interlinked Electronic Register.

### Laboratory data (VL and CD4 count)

Between 2015-2022, there were 2,040,962 VL tests in TIER.Net and 2,179,870 VL tests conducted by NHLS. TIER.Net VL tests increased from 126,205 in 2015 to 352,833 in 2022 while NHLS VL tests increased from 158,648 in 2015 to 358,741 in 2022 (Fig 3A). In 2015, the median (IQR) clinic-level proportion of NHLS VLs captured in TIER.Net was 87.60% (74.52 to 99.60%) for VLs < 50 copies/mL, 84.21% (70.23 to 100.85%) for VLs 50-999 copies/mL, 67.40% (50.58 to 77.50%) for VLs ≥ 1000 copies/mL, and 85.70% (69.97 to 97.85%) overall. By 2022, these had increased to 100.40% (95.75 to 105.14%) for VLs < 50 copies/mL, 92.90% (86.04 to 97.13%) for VLs 50-999 copies/mL, 91.19% (84.65 to 96.58%) for VLs ≥ 1000 copies/mL, and 99.05% (94.53 to 102.53%) overall (Fig 3B). The LMM estimated an annual increase of 1.78% (95% CI: 1.44 to 2.12%) in the proportion of NHLS VLs captured in TIER.Net. In the sensitivity analysis, we found evidence that trends differed by district, with an annual increase of 1.35% (95% CI: 0.88 to 1.83%) in NHLS VL results captured in TIER.Net in eThekwini, and annual increases of 2.50% (95% CI: 1.93 to 3.07%) and 1.48% (95% CI: 0.53 to 2.44%) in uMgungudlovu and uMkhanyakude, respectively (Table 1).

**Table 1:** Estimate (95% confidence interval) of annual change in proportion of NHLS viral load and CD4 count test results captured in TIER.Net between 2015 and 2022, based on linear mixed modelling.

| District | VL estimate (95% CI) | CD4 count estimate (95% CI) |
| --- | --- | --- |
| eThekweni | 1.35% (0.88%,1.83%) | -1.21% (-1.87%, -0.56%) |
| uMgungudlovu | 2.50% (1.93%, 3.07%) | 1.01% (0.23%, 1.79%) |
| uMkhanyakude | 1.48% (0.53%, 2.44%) | 1.46% (0.15%, 2.78%) |

**Fig 3.**
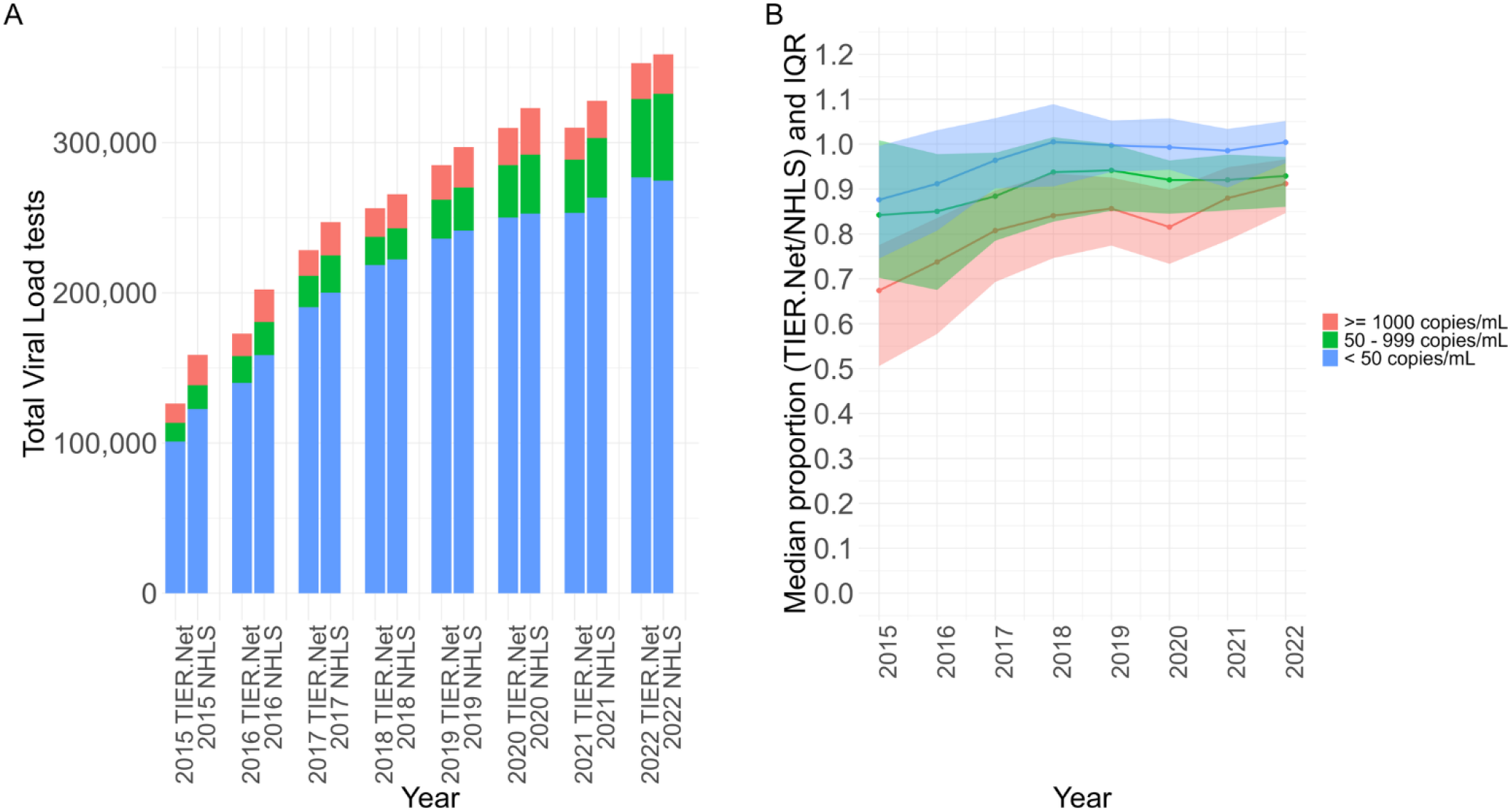
(A) Total number of viral load tests recorded in TIER.Net and NHLS for each threshold by year. (B) Median clinic-level proportion of VLs (TIER.Net/NHLS) with IQR shading for each threshold by year. IQR: interquartile range; National Health Laboratory Service (NHLS); TIER.Net: Three Interlinked Electronic Register.

Between 2015-2022, there were 862,813 CD4 tests in TIER.Net and 1,156,822 CD4 tests conducted by NHLS. TIER.Net CD4 tests decreased from 124,077 in 2015 to 81,886 in 2022 while NHLS CD4 tests decreased from 174,214 in 2015 to 104,751 in 2022 (Fig 4A). In 2015, the median (IQR) proportion of NHLS CD4 tests captured in TIER.Net for clinics was 74.03% (62.44 to 84.66%) for CD4 ≥ 200 cells/µL, 81.80% (IQR 68.17 to 90.33%) for CD4 < 200 cells/µL and 74.31% (63.91 to 85.35%) in total. In 2022, the median (IQR) proportion of NHLS CD4 tests captured in TIER.Net for clinics was 79.21% (66.84 to 89.59%) for CD4 ≥ 200 cells/µL, 81.81% (63.80 to 89.14%) for CD4 < 200 cells/µL and 80.11% (68.40 to 89.13%) for total CD4 tests (Fig 4B). There was insufficient evidence from the LMM to suggest that data capturing had changed over time (annual change: -0.08% [95% CI: -0.56 % to 0.40%]).

**Fig 4.**
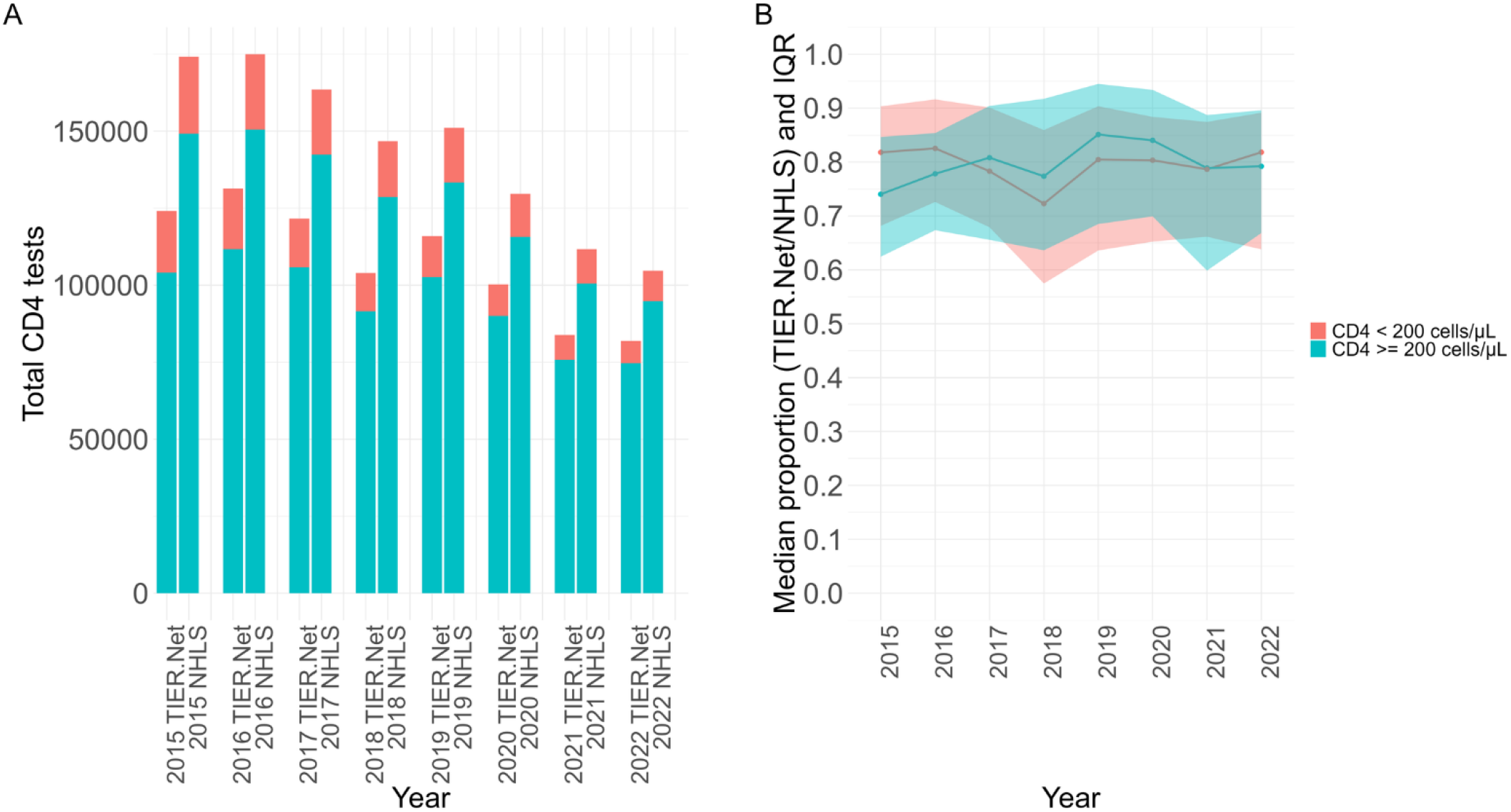
(A) Total CD4 tests from TIER.Net and NHLS for each threshold by year and (B) median proportion (TIER.Net/NHLS) with IQR shading for each threshold for clinics by year. IQR: interquartile range; National Health Laboratory Service (NHLS); TIER.Net: Three Interlinked Electronic Register.

In the sensitivity analysis, we found evidence of a differing trend by district, with an annual decrease of 1.21% (95% CI: -1.87 to -0.56%) in NHLS CD4 results captured in TIER.Net in eThekwini, and annual increases of 1.01% (95% CI: 0.23 to 1.79%) and 1.46% (95% CI: 0.15 to 2.78%) in uMgungudlovu and uMkhanyakude, respectively (Table 1).

## Discussion

We examined the completeness of CCMDD referrals, VL and CD4 count data captured in TIER.Net compared to SyNCH and NHLS over multiple years. Our findings showed that data capture of CCMDD referrals and VLs improved over time, whereas CD4 data capture remained sub-optimal.

TIER.Net CCMDD ART referrals were over-captured in comparison to SyNCH CCMDD ART prescriptions, however this proportion decreased with time. For VL, we found that TIER.Net VL results were under-captured in comparison to NHLS VL results, although the degree of under-capture improved over time. When disaggregating data by VL threshold, we noted that VL < 50 copies/mL had the highest median proportion of NHLS VL captured in TIER.Net, followed by 50-999 copies/mL and then VL ≥ 1000 copies/mL, although IQRs overlapped for most years. The VL tests included in the VL ≥ 1000 copies/mL threshold likely included more repeat VLs which appear out of the annual VL date, and for this reason may be considered lower priority for data capture. Lastly, our analysis showed that TIER.Net CD4 count results were under-captured in comparison to NHLS CD4 count results, and that the degree of under-capture was similar across the CD4 thresholds, CD4 < 200 cells/µL and CD4 ≥ 200 cells/µL. Our sensitivity analysis suggested that the trends in CD4 count capture vary amongst districts with eThekwini observed to have a downward trend, while uMgungudlovu and uMkhanyakude are observed to have upward trends. While CD4 count is a crucial indicator of AHD, there may be less of an emphasis on CD4 counts post-UTT.

There are few other published studies assessing data quality in TIER.Net compared to national electronic information systems on a large scale, and none looking at the quality of decentralised ART delivery data. Our findings on CD4 count data align with earlier research examining first CD4 counts in TIER.Net and NHLS separately between 2004 and 2018 [18]. This study found that fewer first CD4 counts were recorded in TIER.Net compared to NHLS. After UTT implementation, the number of individuals with a first CD4 count declined in TIER.Net and NHLS, however, the decline in NHLS was lower than the decline in TIER.Net. It was suggested that this decline may reflect increased underreporting of laboratory data in TIER.Net [18].

Previous analyses based on comparison to verified records have noted further limitations in the data quality of TIER.Net. A study evaluating the accuracy of patient treatment outcomes (still in care, transferred out, LTFU and deceased) recorded in TIER.Net through comparison with verified records with ART start dates between 2014 to 2017, found that 36% of TIER.Net patient outcomes were misclassified with first CD4 counts amongst other indicators associated with misclassification [19]. Another assessment of TIER.Net data quality used 277 records from October 2018 to December 2019 for HIV and tuberculosis (TB) from three South African districts highlighted areas of improvement for data completeness in TIER.Net [20].

Our study offers a novel contribution by evaluating the completeness of TIER.Net data through comparison with SyNCH for CCMDD data and NHLS for VL and CD4 count data using more recent data. This analysis spans multiple programmatic areas and uses two national-level reference systems to assess data quality across several clinics. The integrated approach provides an understanding of the reliability of routine health information systems used in South Africa’s HIV programme and highlights specific areas where data capture can be strengthened. The limitations of the study were that we included all CD4 count tests, instead of focusing specifically on the first CD4 count at ART initiation which is of higher clinical priority and we had not performed patient-level linkage between TIER.Net and the national reference systems. Additionally, ART regimen information was not available for validation between TIER.Net and SyNCH. Furthermore, under-captured results (VL and CD4 count) in TIER.Net did not confirm if these results were reviewed and intervened at a patient-level by the clinical team.

Strengthening data quality is essential for accurate monitoring of the HIV programme on a national level. TIER.Net is observed to be increasingly reliable for CCMDD and VL data over time, however CD4 counts are under-represented. CD4 counts remain the best indicator of AHD and the implication of under-capturing of CD4 counts, especially <200 cells/µL is that cases of AHD may continue undetected based on TIER.Net data. This gap in data used for monitoring the effectiveness of the HIV programme may underestimate the burden of AHD in KwaZulu-Natal. Recommended strategies include reviewing data workflows and integrating laboratory results into TIER.Net.

In conclusion, our findings indicate progressive improvements in data capture for certain indicators, while highlighting areas requiring further attention. TIER.Net is sufficiently reliable to guide HIV programmes based on CCMDD and VL data. Further research is needed to quantify the accuracy of TIER.Net data following the changes in HIV programme funding in South Africa in 2025.

## Data Availability

The data used for this analysis cannot be shared publicly because of legal and ethical requirements regarding use of routinely collected clinical data in South Africa. The analysis code is available on request from the first author.

## Abbreviations

(ART): Antiretroviral therapy
(CCMDD): Centralized Chronic Medicines Dispensing and Distribution
HIV: (human immunodeficiency virus)
(IQR): Interquartile range
(NHLS): National Health Laboratory Service
(SyNCH): Synchronised National Communication in Health
(TIER.Net): Three Interlinked Electronic Register
(VL): Viral load

## Authorship contributions

JD, NG and LL conceptualised the study. JvdM, LN, VJ and TK oversaw data curation. LN analysed the data, with inputs on the design and implementation from JD and LL. YS, LH, TK, KT, NG and JD were responsible for various components of project administration. LN drafted the manuscript. All authors contributed to interpretation of results, critically reviewed and edited the manuscript, and consented to final publication.

## Acknowledgements

We thank the eThekwini Municipality and uMgungundlovu District Municipality Health Units as well as the staff and patients at the participating healthcare facilities.

